# Prevalence and Associated Factors of Hearing impairment among Children attending Birhanzare Primary School, Addis Ababa, Ethiopia

**DOI:** 10.1101/2023.04.11.23288410

**Authors:** Bitseat W. Haile, Yibeltal M. Feyissa, Betelehem B Kassa, Eyob Kebede Etissa, Haregewoin Asrat, Michael A. Tadesse, Amha Mekasha

## Abstract

**Background:** Hearing impairment is the most prevalent sensory disability globally and a condition of growing concern with around 5.3% of the population having disabling hearing loss. It has significant impacts on the individual and society. The problem is even greater for children as hearing is the main source for language, speech, and cognitive developments.

**Methods:** A school based cross-sectional study at an elementary school in Addis Ababa, Ethiopia was conducted in May 2018. One hundred and ten students from grades 1-5 were enrolled in the study. A structured questionnaire filled by parents/caregivers. Audiometric testing and otoscopic examinations were used to determine hearing impairment and abnormal otoscopic findings respectively. Data was analyzed using SPSS version 25. Descriptive and basic statistical analysis was applied. Logistic regression was used to identify risk factors for hearing impairment. Statistical significance was declared at p values < 0.05.

**Results:** A total of 106 students (27.4% male and 72.6% female) were enrolled in the study. Abnormal otoscopic findings were observed among 30.2% of the participants, with wax being the commonest abnormality observed in twenty-two point eight (22.8%), pus discharge and dull tympanic membrane in 4%, foreign bodies were found in 3 of the participating students (2.8%). The prevalence of disabling hearing impairment in this study (>25db on the better hearing ear), was 34% where the majority (32.1%) had mild hearing impairment. Abnormal otoscopic finding showed significant association with hearing impairment.

**Conclusion:** The prevalence of hearing impairment in our study is higher than WHO and other regional estimates. Given the high prevalence of HI, availability of management options for mild HI, and its underlying causes, early hearing screening programs in children should be integrated into existing systems such as the Ethiopian school health program as well as early childhood growth and development monitoring entry points.

## Background

Sensory experiences dictate how we interact with our environment. Regardless of age normal hearing affects individual’s psychological development. It is also essential for the development of auditory abilities which in turn has profound impact on oral language acquisition, verbal comprehension, reading and writing acquisition as well as learning and academic performance (Etges et al., 2012; Guerra-Silva, 2012; Shoukrya, 2010). Despite hearing impairment (HI) being an issue at all ages, the impact is more pronounced in children where the effect extends to language, speech, and cognitive abilities as well as social and self-image domains.

Otitis media, one of the most common childhood diseases, is a leading cause of hearing impairment in children (Ross et al., 2008; Skarzyński et al., 2016; Taha et al., 2010). Listening to loud music has also become an increasing cause of high frequency HI in this group (Chan et al., 2020; Mahboubi et al., 2013). Other notable causes of HI in school aged children include other infectious diseases (mumps, measles, and meningitis) as well as mechanical injuries (Nelson et al., 2002; Yamatodani et al., 2013).

The learning impacts from HI mostly present in the first year of school, while many children with mild show considerable learning difficulties in the third grade. This has been attributed to earlier losses in learning ability, changes in language complexity, loss of visual cues and required auditory capacity (Sousa, 2009).

Early diagnosis of HI allows detection and referral to specialists for care providing opportunities to salvage children’s cognitive, social, emotional, and communicative alterations. Choosing the strategy and setting for early screening interventions should consider the populations health seeking ability, access to services, socio-cultural considerations as well as parental know how and cooperation. The later, has been challenging in many settings with governments/providers opting for school-based schemes where majority of children gather in academic centers and can all be examined. (Ref) Hearing loss or impairment have consistently ranked the most prevalent sensory disability globally for many decades. According to the latest data from WHO deafness and hearing loss, 360 million people (5.3%) of the world’s population had disabling hearing loss defined as hearing loss greater than 40dB and 30dB in the better hearing ear in adults and children, respectively (World Health Organization & World Health Organization, 2018).

The burden affects older adults, the pediatric group and particularly the Sub-Saharan Africa population: one in three adults 65years or older, nine out of 100 children affected (32 million), where SSA contributed to a much larger share in the pediatric group, at nine percent (WHO, 2021, n.d.): the true prevalence may even be higher than WHO estimates. A systematic review investigating the prevalence of HI is SSA in school and population settings found a much higher prevalence. Using WHO cutoff of 30dB for disabling HI the prevalence was found to be 6.6% and 17% for school and population-based studies, respectively. The prevalence for mild HI with a cutoff point of 25dB in the same study (median) was 7.7% and 17% in similar study settings (Mulwafu et al., 2016). Several studies have been conducted to measure the prevalence, magnitude, risk factors and associations of HI in children.

*S*everal studies have been reviewed on the prevalence of HI in many African nations with varying findings. Some studies reported HI prevalence lower than or closer to WHO estimates (9% in Sierra Leone school children, 7·5% in school children in urban areas in SA, and 4.1 % in Swaziland (Minja & Machemba, 1996; Skarzyński et al., 2016; Taha et al., 2010), two studies from Tanzania and Egypt showed a higher prevalence. The study from Tanzania showed a higher prevalence (14.1% Vs 7.7%) in school children in rural area with chronic suppurative otitis media being linked to the increased prevalence in rural areas (Minja & Machemba, 1996). There are not many large populations and school-based studies conducted in Ethiopia. The largest study on the national prevalence of HI in Ethiopia comes from a cross-sectional, community-based study among households under demographic and health surveillance in eastern Ethiopian children aged 0– 14years.from a total of 21,572 children screened for disability 586 (2.7%) had at least one kind of disability, hearing impairment being the most reported disability; 417(71.2%) (Geda et al., 2016). A community-based study from Kersa, Eastern Hararge, Ethiopia confirms HI as a leading cause of parent reported disability with 41% of children with HI having chronic ear discharge.

Most existing hearing screening programs in LMICs stem from local initiatives supported by Western countries, while China and few Eastern European countries have included it as part of bigger national programs (Wen et al., 2022). Many hearing Screening Programs have demonstrated their relevance as the primary means for early detection of hearing loss in children. Up until now, the programs performed in schools have been offered as local initiatives in places such as the United States, Australia, China, and a few European countries.

The Ethiopian Federal Ministry of Health has integrated hearing screening as part of the school health program among the 10 health service packages to be introduced to schools from nursery to tertiary institutions. This study aims to provide a platform for assessment of hearing impairment in schools as part of routine health care provision for children attending schools. It will also serve as a base line for further population-based research.

Thus, this study attempts to assess the prevalence and associated factors of hearing impairment in school children attending Birhaneh Zare school, Addis Ababa, Ethiopia.

## METHODOLOGY

### Study setting

A school based cross-sectional study was conducted in May 2018 at Birhaneh Zare Primary school, Addis Ababa, Ethiopia. From the 10 sub cities in the capital lottery method was applied where Bole sub city and subsequently the Woreda 3 where Birhaneh Zare, the only primary governmental school, is situated was selected.

### Sampling

A single population proportion formula, a 10% non-response rate and estimated HI prevalence of 7.7% (Mulwafu et al., 2016) was used to determine the sample size for the study, 120. From the 322 students enrolled in grades 1-5, 120 were selected for the study through simple random sampling of students from the schools’ registry. Due to lack of parental consent in five and lack of assent in other five 110 students were evaluated, of which four were absent from school at the time of the evaluation and screening. Since the procedure requires the students to comprehend and reply to instructions of the screening preschool students were excluded.

To assess the prevalence of hearing impairment students from grades 1 through 5 was included. Of the 322 students attending grades 1 through 5, 120 were taken by quota from each class. Students were evaluated in a five-day screening program which was held on tea and lunch break hours; so, students did not miss any class.

### Sampling strategy

Figure 1 below shows steps in the sampling strategy.

**Figure 1:**
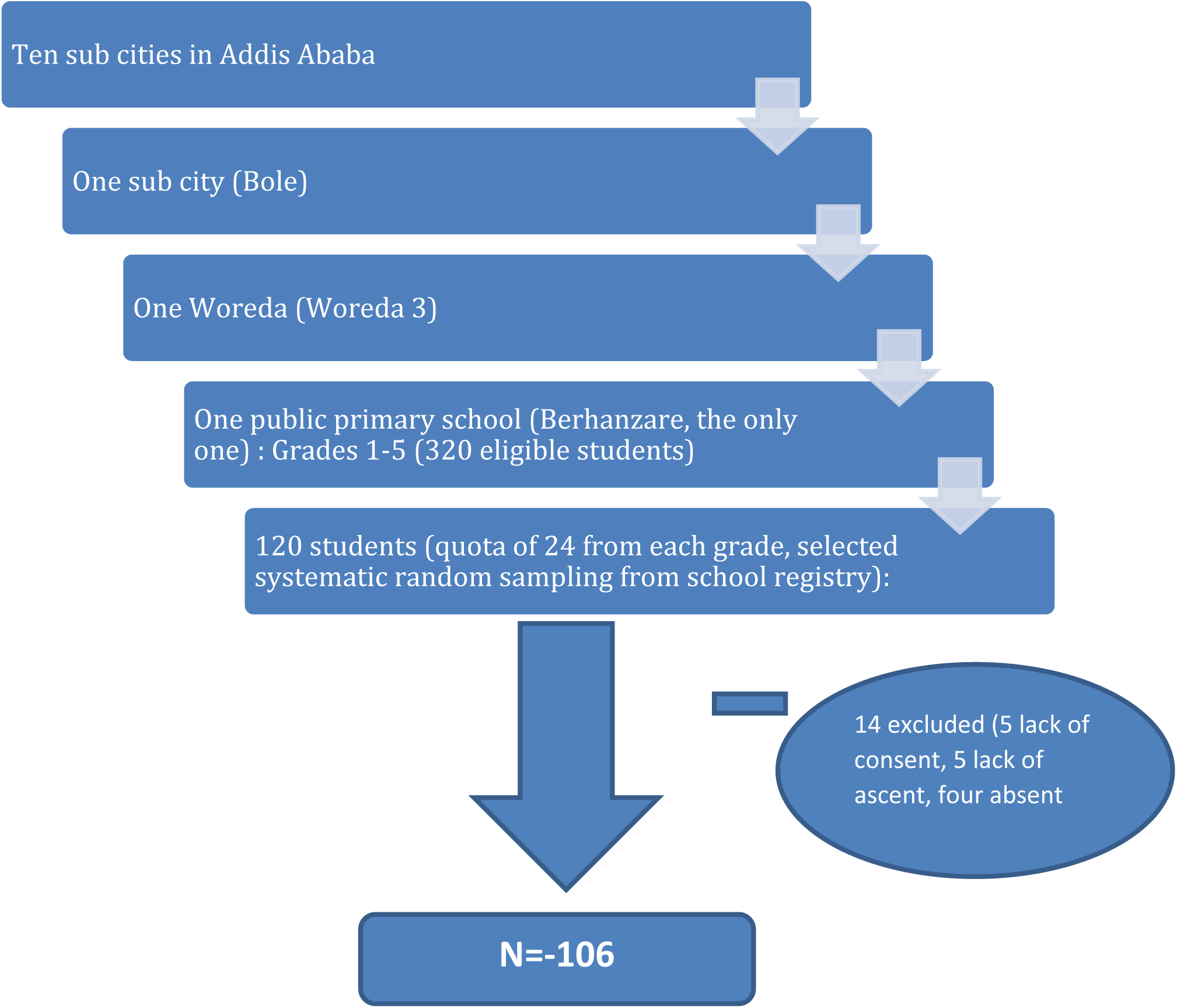
Flow gram sampling strategy

#### Inclusion and exclusion criteria

Inclusion criteria

- All Children grades 1 through 5 attending the school whose parents or guardians give consent and the students give verbal consent.

Exclusion criteria

- Students who are not willing to participate in the study or lack of consent from parents or guardians were excluded.
- Students who fail to obtain written consent from parents or guardians.

### Data collection

Data was collected by trained audio metrists, in a quiet room; using a screening Audiometer AS 208 was used where hearing was assessed at frequencies of 500Hz, 1000Hz and 2000HZ. The students were oriented of the procedure to raise their hand every time they hear a beep even if very soft and to put hands down once the beep goes away; headphones were applied. And the child’s hearing status was recorded and compiled for analysis (see Annex3). If student responds to all tones in both ears; marked as pass. But if student fails to respond to any of the tones, marked fail and refer for further screening.

To evaluate risk factors and associations, a structured questionnaire (developed in English, translated to Amharic version, and tested through back translation) was availed for parents to fill at home and send back. This was then compiled and added to a child’s hearing test for further analysis on association for hearing condition. Otoscopic evaluations were performed by two ENT residents.

### Bias

Two biases were expected in this study, the healthy child effect and social desirability bias. Rigorous testing and reverse translation of patient information about participating in the study, where any relevant findings would be availed for use for participants in the study in their future enrollment into care was applied to address the later bias, while the healthy child effect minimized through an extended five days period of data collection.

### Variables

#### Dependent variable

- Hearing impairment

#### Independent variables

- Age, Sex, History of Neonatal admission. History of Treatment for meningitis, History of head injury, Gestational age at birth (term/preterm), Recurrent ear infections, Family member with hearing impairment.

### Data quality assurance

Audio metrist was oriented on specific procedure to be used by the principal investigator on measurements and classifications of hearing impairment. The data collection format of the data collector was checked daily for completeness by the principal investigator. Data collectors were also trained on the study sample collection including recruitment of participants, how to fill out the data collecting instrument and ethical issues such as confidentiality.

### Data analysis

Data was compiled and analyzed using SPSS version 25 software package. Bivariate analysis was used and variables with P-value of 0.3 or less were analyzed using logistic regression methods. Odds ratio with 95% CI generated using logistic regression was employed to determine independent factor associated with hearing impairment. In the final multivariable binary logistic regression model, after checking for model-fitting information (Hosmer and Lemeshow test p=0.356). Results were reported as statistically significant whenever P value was less than 0.05

### Ethical Considerations

Following standard procedure, ethical approval was granted by Depratment of Pediatrics Ethics and research Committee, at the College of Health Sciences of the Addis Ababa University, Ref.No-Pd/Mf/329/23. Formal consent was obtained from all parents/guardians for participation in the study using written consent forms in local language.

## RESULTS

### Demographic profile

A total of 106 students participated in this study. Among the participants 77 (72.6%) were female and 29 (27.4%) were male. The age distribution of the students showed a normal distribution pattern on normality curve with skewedness. The mean age (SD) was 10 (2) years. The minimum observed age of the participants was 7 years whereas maximum was 16 years. The gestational age of the respondents at the time of delivery was assessed. Accordingly, 8 (7.5%) were preterm and98 (84%) were delivered at term. History of hospital admission as a neonate was elicited among 11 (10.4%) of the participants. (Table 1)

**Table 1:**
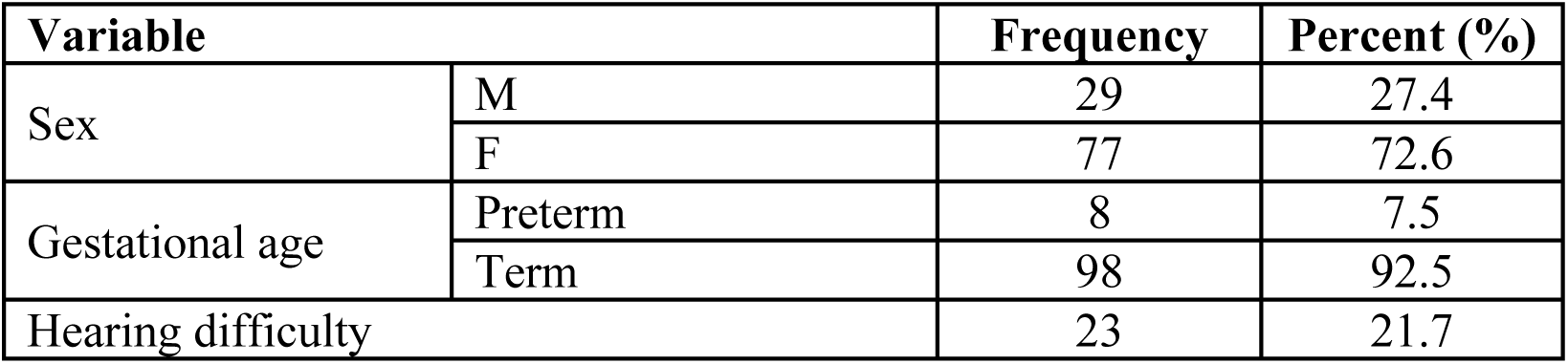
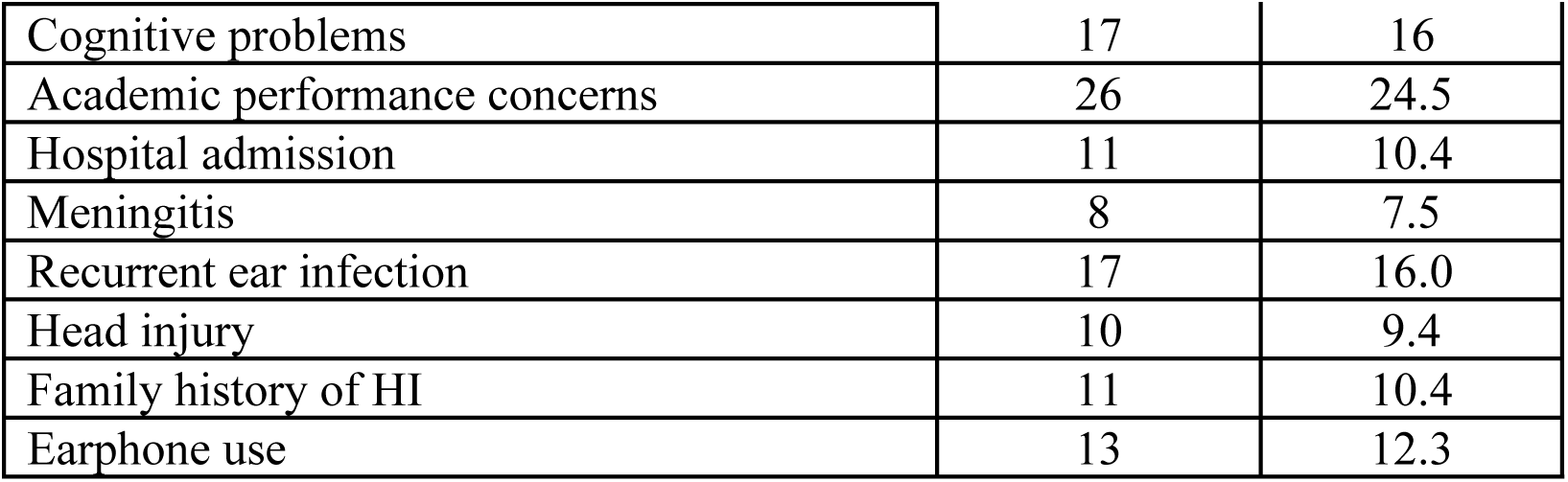
Baseline characteristics

| Variable |  | Frequency | Percent (%) |
| --- | --- | --- | --- |
| Sex | M | 29 | 27.4 |
|  | F | 77 | 72.6 |
| Gestational age | Preterm | 8 | 7.5 |
|  | Term | 98 | 92.5 |
| Hearing difficulty |  | 23 | 21.7 |

|  |  |  |
| --- | --- | --- |
| Cognitive problems | 17 | 16 |
| Academic performance concerns | 26 | 24.5 |
| Hospital admission | 11 | 10.4 |
| Meningitis | 8 | 7.5 |
| Recurrent ear infection | 17 | 16.0 |
| Head injury | 10 | 9.4 |
| Family history of HI | 11 | 10.4 |
| Earphone use | 13 | 12.3 |

Worry about the child’s hearing ability was reported among 23 (21.7%) of the respondents, complaint about intellectual capacity of the child was reported among 17 (16%) of the respondents. Parental worry about the academic performance of the child was mentioned among 26 (24.5%) of the participants. Some associated risk factors for hearing impairment were explored. History of meningitis was elicited among 8 (7.5%) of children; history of recurrent ear infection was reported among 17 (16.0%) of the children. Family history of hearing difficulty was present in 11 (10.4%) of the cases. History of previous head injury was elicited in 10 (9.4%) and frequent use of earphone was reported in 13 (12.3%) of the cases. (Table 1)

### Otoscopic findings

Otoscopic examination of the participants showed abnormal finding among 30.2%. Of the abnormal findings, wax was the commonest abnormality observed in 22.8% of the cases, pus discharge and dull tympanic membrane was observed in 4% of the cases, tympanic membrane perforation and foreign body was detected among 3% and 2% of children, respectively. Tables 2 and 3 show the audiometric and otoscopic findings respectively.

**Table 2:** Otoscopic Findings

| Test Results | Frequency | Percentage |
| --- | --- | --- |
| Wax | 23 | 22.8 |
| Perforation | 3 | 3 |
| Other | 6 | 6 |

**Table 3:** Audiometric Findings

| Test Results | Frequency | Percentage |
| --- | --- | --- |
| <26db | 70 | 66.0 |
| 26-40db | 34 | 32.1 |
| >40db | 2 | 1.9 |

**Table 4:** Bivariable and multivariable results of predictors of hearing impairment.

| Variable |  | COR<br>(95% CI) | P-value | AOR<br>(95% CI) | P-value |
| --- | --- | --- | --- | --- | --- |
| Sex | Male | 0.52 (0.20-1.38) | 0.194 | 0.49 (1.17-1.41) | 0.190 |
|  | Female | 1 |  | 1 |  |
| Gestational age | Pre-term | 1.18 (0.26-5.25) | 0.826 |  |  |
|  | Term | 1 |  |  |  |
| Hearing difficulty | Yes | 1.68 (0.65-4.43) | 0.279 | 1.69 (0.52-5.42) | 0.376 |
|  | No | 1 |  | 1 |  |
| Cognitive problem | Yes | 1.07 (0.36-3.18) | 0.899 |  |  |
|  | No | 1 |  |  |  |
| Academic problem | Yes | 1.61 (0.64-4.01) | 0.303 |  |  |
|  | No | 1 |  |  |  |
| Hospital admission | Yes | 1.12 (0.31-4.12) | 0.859 |  |  |
|  | No | 1 |  |  |  |
| Meningitis | Yes | 2.06 (0.48-8.78) | 0.327 |  |  |
|  | No | 1 |  |  |  |
| Otitis media | Yes | 3.87 (0.13-1.11) | 0.078 | 2.81 (0.81-9.79) | 0.103 |
|  | No | 1 |  | 1 |  |
| Head Injury | Yes | 1.33 (0.35-5.06) | 0.673 |  |  |
|  | No | 1 |  |  |  |
| Family history | Yes | 1.12 (0.31-4.13) | 0.859 |  |  |
|  | No | 1 |  |  |  |
| Earphone use | Yes | 1.25 (0.37-4.14) | 0.715 |  |  |
|  | No | 1 |  |  |  |
| Otoscopy | Normal | 1 |  | 1 |  |
|  | Abnormal | 3.28 (1.37-7.81) | 0.007 | 4.05 (1.59-10.34) | **0.003** |

### Audiometric findings

Assessment for hearing impairment was done objectively by using screening pure tone audiometer. A cut off point of 25db on the better hearing ear was used to define disabling hearing impairment as set by WHO standard guideline. The prevalence of disabling hearing impairment in this study, that is ≥ 26db on the better hearing ear, was 34.0%. Of which 32.1% had mild hearing impairment and moderate to severe impairment was present among 2% of them. No child had profound hearing impairment in this study. (Table 3)

### Predictor of hearing impairment

Variables from the bivariate analysis with a p-value <0.3 were taken into the final model. On Bivariate analysis the variable observed to have statistically significant association were presence of abnormal otoscopic finding (COR = 3.28, 95% CI 1.37, 7.81, p-value = 0.007). In the multivariable binary logistic regression after adjusting for covariates only otoscopic finding remains to be significantly associated with hearing impairment. Students with abnormal otoscopic finding had 4 times increased risk of hearing impairment compared to those with normal otoscopic finding (AOR = 4.05, 95% CI 1.59, 10.34, p-value = 0.003).

## DISCUSSION

Reported prevalence of hearing impairment is variable in different geographic areas. Rural children are more affected than their urban counterparts. In our study the prevalence of hearing impairment was 34% when >25db was used as a cut-off point to diagnose hearing impairment. The prevalence was 17.6% when a 30db is used as cut off point. Prevalence was variable in different studies from developing countries. But similar results were observed from Egypt, 20.9%, Tanzania, and Tajikistan 24% (Mulwafu et al., 2016; Shoukrya, 2010; Skarzyński et al., 2016). The heterogeneity of prevalence among studies may be due to difference in cut-off point used, whether the study is done in rural or urban setting, and other factors.

When we assessed the severity of HI, 32.1% of them were mild HI with moderate to severe impairment accounting for the rest of 2% of HI. A similar finding was reported from a study in Egypt where no child had profound hearing impairment (Taha et al., 2010). This could be because profound hearing impairment wouldn’t allow students to attend schools.

It has been reported that the negative consequences of HI can occur regardless of severity of the HI although more severe and more prolonged HI may have more consequences. Even minimal HI was associated with higher stress, low self-esteem, and a higher rate of failure at school (Taha et al., 2010). Therefore, even minimal, unilateral, and fluctuating HI should be considered significant and appropriate interventions should be made timely. As the prevalence of HI was high and found at early stages, the impact can be mitigated through early screening and enrollment into care.

Different factors can be associated with higher risk of HI. In our study among factors assessed in this regard, presence of ear wax and ear infection strongly correlated with HI. Of the children with ear wax, 43.5% of them had HI. This finding is comparable to other studies reported from Brazil, India, and Egypt. Even though HI can resolve with removal of impacted cerumen, it was a predictor of permanent hearing impairment and was as well associated with low educational achievement in children. Of those with otoscopic of untreated ear infection 57.1% had hearing impairment which shows 4 times increased risk than those students with no evidence of ear infection. Similar results were observed in a Sierra Leonean and Tanzanian study showing untreated ear infections as cause of hearing impairment (Cai & McPherson, 2017; Mulwafu et al., 2016).

### Limitation of the study

The study was conducted in a single school with small sample size. There was recall bias by parents/ guardians to the perinatal history.

## CONCLUSIONS

The prevalence of HI among students attending Birhaneh Zare primary school A.A. is 34%. Majority of the participants of the study had mild HI. Wax was the most common identified underlying risk factor and association was found between hearing impairment and otoscopic finding of ear infection.

The prevalence of HI in our study is high and screening programs in children aimed at early detection and interventions should be formulated and screening audiometers utilized by the ministry of health in cooperation with ministry of education.

A larger scale school based as well as community-based studies should be conducted to assess prevalence of HI in different areas and regions of the country

## Data Availability

all data will be provided

## Notes

### Competing Interest Statement

The authors have declared no competing interest.

### Funding Statement

this research was done as partial fulfillment for pediatric specialty certificate training programme.as per the school's regulation funding was provided by Addis Ababa University College of Health Science

### Author Declarations

Addis Ababa University college of health sciences department of pediatrics ethics and research committee gave the ethical clearance through the regular procedure, Ref.No-Pd/Mf/329/23

